# The cerebellum and cognitive function: anatomical evidence from a transdiagnostic sample

**DOI:** 10.1101/2023.02.22.23286149

**Authors:** Indrit Bègue, Yannis Elandaloussi, Farnaz Delavari, Hengyi Cao, Alexandra Moussa-Tooks, Mathilde Roser, Pierrick Coupé, Marion Leboyer, Stefan Kaiser, Josselin Houenou, Roscoe Brady, Charles Laidi

**Author notes:** **Corresponding author** Indrit Bègue MSc PhD FMH, Department of psychiatry, Beth Israel Deaconess Medical School & Harvard Medical School; Department of psychiatry, McLean Hospital & Harvard Medical School, Boston, Massachusetts, USA.

## Abstract

**Introduction:** The cerebellum, most known for its role in motor control, exerts a key role in cognition. Multiple lines of evidence across human functional, lesion and animal data point to a role of the cerebellum, in particular of Crus I, Crus II and Lobule VIIB, in cognitive function. However, whether cerebellar substrates pertaining to distinct facets of cognitive function exist is not known.

**Methods:** We analyzed structural neuroimaging data from the Healthy Brain Network (HBN). Cerebellar parcellation was performed via a standard validated automated segmentation pipeline (CERES) with stringent visual quality check (n = 662 subjects retained from initial n = 1452). We used data-driven canonical correlation analyses (CCA) to examine regional gray matter volumetric (GMV) differences in association to cognitive function assessed with the NIH Toolbox Cognition Domain (NIH-TB). Our multivariate analyses accounted for psychopathology severity, age, sex, scan location and intracranial volume.

**Results:** Multivariate CCA uncovered a significant correlation between two components entailing a latent cognitive canonical variate composed of NIH-TB subscales and the brain canonical variate (cerebellar regions’ GMV and intracranial volume, ICV). A bootstrapping and a permutation procedure ensured the results are statistically significant and the CCA model, stable. The identified components correspond to only partly shared cerebellar -cognitive function relationship with a first map encompassing cognitive flexibility (r=0.89) and speed of processing (r=0.65) associated with regional gray matter volume in Crus II (r=0.57) and Lobule X (r=0.59) and a second map including the Crus I (r=0.49) and Lobule VI (r=0.49) associated with cognitive control (r=-0.51). Working memory associations were similarly present in both these maps (Crus II, Lobule X, Crus I and Lobule VI) for the first (r=0.52) and second (r=0.51) component.

**Discussion:** Our results show evidence in favor of structural sub-specialization in the cerebellum, independently of psychopathology contributions to cognitive function and brain structure. Overall, these findings highlight a prominent role for the human cerebellum in cognitive function for flexible and stable adaptive behavior.

## Introduction

The cerebellum is a fascinating infratentorial brain structure with a pivotal role in human cognition (Schmahmann et al. 2019). Research on the cerebellum has been traditionally limited to its role in motor control, even though the majority of the cerebellar cortex is not involved in motor action planning or execution (King et al. 2019). Cerebellar lesions across different diagnostic entities are associated with a diverse palette of cognitive deficits including disturbances of executive function such as planning, set-shifting, working memory and verbal fluency (Schmahmann and Sherman 1998).

A recent study linked cerebellar anatomy to cognitive functioning and found that anatomical features predicted both *general* cognitive function and psychopathology (Moberget et al. 2019). However, regional cerebellar morphometry differences relating to *general* cognitive function in psychosis (Moussa-Tooks et al. 2022) or in autism (Laidi et al. 2022) were no different from controls. One reason for the discrepancy could be the difference in these studies’ approaches (dimensional vs. case-control comparisons). These studies looked into *general* cognitive function which may fail to capture anatomy-cognition links in the cerebellum because the relationships possibly concern specific cognitive domains. Studies are starting to progressively elucidate the functional organization of the cerebellum (King et al. 2019; Buckner et al. 2011; Guell et al. 2018). Yet, a finer-grained nuanced investigation of the distinct facets of cognition is currently lacking leaving open the question of whether a structural cerebellar subspecialization exists with respect to cognitive abilities. Previous evidence showed that brain lesions in Crus I and Crus II, VIIB (Stoodley et al. 2016; Stoodley and Schmahmann 2010; Kansal et al. 2017) and to a lower extent VIIIA and VI are associated with executive function performance. A seminal study in patients with cerebellar degeneration showed that distinct components of cognitive function (e.g. executive function, working memory, perceptual processing and so on) relate differently to cerebellar topography (Kansal et al. 2017). Nevertheless, even though lesion studies are informative they have limitations, and a large-scale examination of the cerebellar mapping of distinct cognitive components is lacking. Furthermore, it is worth noting that psychopathology severity (e.g., levels of anxiety, depression and so on) has not been systematically accounted for in the reviewed studies examining associations with cognitive function. Psychopathology severity impacts cognitive function (Chavez-Baldini et al. 2021) but also brain structure properties particularly in the developing brain (Patel et al. 2022; Mattoni, Wilson, and Olino 2021; Romer, Ren, and Pizzagalli 2023). Recently, cerebellar structure has been shown to be linked to both general cognitive function and psychopathology (Moberget et al. 2019). However, how cognitive mapping in cerebellar anatomy is represented independently of psychopathology contributions is not fully elucidated. Such investigation is of key interest with respect to the ensuing potential for clinical (e.g. neuromodulation) translation (Yao et al. 2022). In sum, a major gap remains in the current understanding of cerebellar contributions to cognitive function and psychopathology: it is not clear whether sub-specializations in cerebellar anatomy pertaining to components of distinct cognitive function exist and whether such differences can be observed independently of psychopathology severity.

In the current investigation, we examined for the first time how cerebellar regional anatomy may support cognitive function, capitalizing on a large dataset of transdiagnostic population employing a dimensional approach in agreement with the Research Domain Criteria framework (Cuthbert 2014). Our aim was to first outline gray matter volume changes in the cerebellum across distinct facets of cognitive function (e.g., executive function, working memory, cognitive flexibility, processing speed). Considering we were interested in cohorts with detailed cognitive phenotyping, we used the Healthy Brain Network (HBN) (Alexander et al. 2017), a landmark transdiagnostic mental health neuroimaging and behavioral dataset in a few thousand children and adolescents. The HBN includes predominantly unmedicated children and teenagers allowing us to examine cognitive cerebellar correlates unconfounded by chronic psychotropic consumption. Because spurious results can arise from quality control issues regarding neuroimaging scans, a rigorous quality assessment with visual inspection of all images is key to ensure the robustness of the results (Laidi et al. 2022). We used a data-driven multivariate canonical correlation analysis model (CCA) to evaluate the association between cerebellar anatomy and cognitive phenotype. Importantly, we used both permutation testing and bootstrapping to assess the significance and the robustness of our results.

## Methods

### Subjects

In the current investigation, we used data coming from an openly shared dataset, the Healthy Brain Network (HBN) project (Alexander et al. 2017). The HBN is a transdiagnostic dataset of neuroimaging and psychopathological assessments from a cohort of psychiatric or at-risk population of children and adolescents (5-21 years) (Alexander et al. 2017). Participants with severe neurological disorder or acute psychotic episodes are excluded in this cohort. In our study, considering our focus was on neurocognitive functioning, we excluded subjects with an intellectual deficiency (age-corrected IQ below 70, as measured with the Wechsler Adult Intelligence Scale (WASI-II) or the Wechsler Intelligence Scale for Children (WISC-V) (Na and Burns 2016). The full clinical assessment of the HBN cohort is described elsewhere in depth (Alexander et al. 2017).

### Assessments

We were motivated to examine distinct contributions of the specific cognitive function aspects to the cerebellar anatomy. In the HBN database, cognition is quantified via the NIH Toolbox Cognition domain (Weintraub et al. 2013). The four subscales of the NIH Toolbox Cognition domain used in the HBN are detailed as follows. 1) The NIH Flanker assesses inhibitory executive control and attention and requires participants to focus on a target stimulus and ignore flanking stimuli. 2) NIH Card Sort assesses cognitive flexibility and requires participants to apply one rule to two target pictures (e.g., matching by color) and then another (e.g., matching by shape). 3) NIH List assesses working memory function and requires participants to sequence visually and orally presented stimuli e.g. by size. 4) NIH Pattern Comparison Processing Speed Test assesses processing speed by requiring participants to compare two side-by-side pictures (same vs. different). In all subscales, higher scores mean better ability. We used the standardized normative scores for each subscale. Psychopathology severity is quantified with the Child Behavior Checklist (CBCL) (Achenbach and Rescorla 2001), a widely employed scale that measures emotional, behavioral and social problems in children and teenagers of 1.5 - 18 years old. These scales have a mean t-score of 50 with a standard deviation of 10. A t-score ≤ 64 indicates non-clinical symptoms, a t-score between 65 and 69 indicates problems rated high enough to be of concern but not overtly deviant, a t-score ≥70 indicates clinical symptoms (Achenbach and Rescorla 2001; Carta et al. 2020).

### MRI acquisition

Acquisition of MRI scans was done in three sites in New York City: Staten Island, Rutgers University and Cornell Brain Imaging Center. The specific details of each acquisition protocol are as follows: Staten Island images were acquired on a 1.5T Siemens Avanto (*TR* = 2730 ms, *TE* = 1.64 ms, flip angle = 7°, slice number= 176, voxel dimensions = 1.0 × 1.0 × 1.0 mm^3^). Rutgers University images were acquired on a 3T Siemens Tim Trio (*TR* = 2500 ms, *TE* = 3.15 ms, flip angle = 8°, slice number= 224, voxel dimensions = 0.8 × 0.8 × 0.8 mm^3^). Cornell Brain Imaging Center images were acquired on a Siemens Prisma 3T MRI (*TR* = 2500 ms, *TE* = 3.15 ms, flip angle = 8°, slice number= 224, voxel dimensions = 0.8 × 0.8 × 0.8 mm^3^)

### MRI processing

All subjects were processed using the CERES pipeline (Romero et al. 2017). This fully automated method relies on an multi-atlas patch-based strategy that has been compared with manual tracing and performs well compared to other segmentation methods (Carass et al. 2018). All structural T1 MRIs were processed by PC on a high computing performance cluster in Bordeaux, France. The CERES pipeline follows the parcellation protocol described in (Park et al. 2014) (), which provides a parcellation of the cerebellum and gray matter volumes for all cerebellar Lobules, with the exception of the cerebellar vermis, which is included in every Lobule. Moreover, the CERES pipeline provides a mask of intracranial volume (ICV) (Manjón et al. 2014).

### Quality control

The quality control procedure was done in two steps: 1) visual inspection of the raw T1 images 2) visual inspection of the images issued from the parcellation procedure in every slice for each spatial plan of the cerebellum by an expert rater (YE) - blind to the clinical features of each participant. We identified subjects with non-cerebellar voxels labeled as voxels belonging to the cerebellum, and vice versa, and subjects with parcellation errors within the cerebellar lobules. The same procedure has been applied previously (Laidi et al. 2022). No images with parcellation defects were included in further analyses. After the preprocessing and quality control step we excluded 602 individuals after visual inspection of the raw T1 images (280 individuals), parcellation errors (279 individuals), low IQ (43 individuals) and incomplete psychometric scores (60 individuals). A final sample of 662 individuals was included in the subsequent neuroimaging analyses. A summary of the repartition of the excluded subjects can be found in supplementary figure S1.

## Statistical analyses

### Canonical correlation analysis

We performed our multivariate analyses with scikit-learn library (Pedregosa et al. 2011). We employed a regularized kernel Canonical Correlation Analysis (CCA), using an open-source python *pyrcca* package (Bilenko and Gallant 2016), as a multivariate approach to evaluate the association between cerebellar anatomy (component with anatomical features “X”) and cognitive phenotype (component with clinical scores of interest “Y”). In brief, CCA solves the canonical spaces in which the maximal correlation of projected datasets occurs, not pre-assuming the directionality of the relationship between datasets (Hardoon et al. 2007). One documented disadvantage of CCA is overfitting to noise correlation of the datasets. To overcome this limitation, we implemented the algorithm used by the *pyrcca* toolbox. This algorithm constrains the number of components and find the optimal regularization parameters in a data-driven manner using a 10-fold cross validation approach (Bilenko and Gallant 2016). Here, we investigated a range of components ([2, 3, 4]) and a range of parameters ([0.0001, 0.01, 1, 100]). Such step resulted in the best number of components of 2 and best regularization parameter of 0.0001 that were used for the subsequent analyses.

In our analysis, the anatomical component included our regions of interest, namely the anterior lobe (Lobules I – V), Lobule VI, Crus I, Crus II, Lobule VIIB, VIIIA, IX and X and the ICV, after regressing out the effect of scan location, age and sex. We included in the cognitive component the standardized scores of the subscales of NIH toolbox, namely List subscale indexing working memory, Card subscale indexing cognitive flexibility, Flanker’s subscale indexing cognitive control and Processing subscale indexing processing speed. We then computed the correlation between the two canonical components for clinical and anatomical features. We repeated these analyses by including psychopathology severity quantified by the total t-score of the Child Behavior Checklist (CBCL) (Mazefsky et al. 2011).

### Assessment of statistical significance and model stability

To assess statistical significance, we used non-parametric permutation testing (Ericson and Zinoviev 2001). Permutation testing involves random rearrangement of samples without replacement in order to estimate the population distribution and in turn, test the null hypothesis. Thus, p-value would be defined as the proportion of permuted samples that test statistically higher than our observed sample. In this study, unless specified otherwise, the threshold of significance was set to p=0.05 corresponding to the r value higher than the r value of the 95 percentile in a 10’ 000 random permutation test.

Crucially, we assessed the model stability through bootstrapping analysis (Carpenter and Bithell 2000). Bootstrapping is used to create a sampling distribution by repeatedly taking random samples with replacement from the original sample. We then performed CCA on each bootstrapped sample and collected these results to perform summary statistics (mean and confidence intervals). The average estimated from these multiple random samples may be used to infer results regarding the robustness of the CCA results for the original sample. Here, we used a bootstrap of 10’000 random samples with replacement which showed a normal distribution of canonical correlations. The results were considered stable if the 95% confidence interval of the bootstrap distribution would not include zero.

## Results

### Study sample: demographics and psychopathology

The demographic characteristics of our sample can be found in Table 1. In sum, the cohort included in our study (n=662), had a mean age of 10.5 years [range 5.82 – 17.74 years old] and were predominantly males (58%).

**Table 1.** Summary of cohort characteristics (age, sex, scan site and intracranial volume) and cognitive characteristics (NIH toolbox subscales) of our cohort (n=850). *Abbreviations*: ICV, intracranial volume; NIH Toolbox = NIH-TB; NIH List=NIH TB List Sorting Working Memory Test; NIH Card= NIH TB Cognition Domain Dimensional Change Card Sort Test; NIH Flanker = NIH TB Flanker Inhibitory Control and Attention Test; NIH Processing = NIH-TB Pattern Comparison Processing Speed Test; CBCL total = total t-score of Child Behavior Checklist. CBIC: Cornell Brain Imaging Center; SI: Staten Island; RU: Rutgers University; ‘M’=Male; ‘F’=Female

| Variable | Count | Mean | Standard Deviation | Min | Max |
| --- | --- | --- | --- | --- | --- |
| Age (in years) | 662 | 10.5 | 2.91 | 5.82 | 17.74 |
| Sex | {'M': 385, 'F': 277} |  |  |  |  |
| ICV (in cm3) | 662 | 1410.59 | 136.57 | 1027 | 1867.81 |
| Scanning site: CBIC | 285 |  |  |  |  |
| Scanning site: RU | 315 |  |  |  |  |
| Scanning site: SI | 62 |  |  |  |  |
| NIH Card | 662 | 93.46 | 15.49 | 53 | 146 |
| NIH Flanker | 662 | 88.61 | 14.01 | 60 | 151 |
| NIH List | 662 | 97.13 | 14.69 | 55 | 169 |
| NIH Processing | 662 | 94.59 | 23.56 | 20 | 169 |
| CBCL Total | 662 | 57.54 | 11.38 | 24.0 | 82.0 |

Next, we investigated the relationship between cognitive function assessed with the NIH-TB subscales and psychopathology severity quantified by the CBCL total t-score. We found positive correlations between the individual cognitive subscales scores with small effect sizes (Figure 2), indicating that these subscales measure little overlapping constructs. However, there were no associations of the cognitive subscales with psychopathology severity (CBCL) indicating no significant impact of psychopathology on distinct components of cognitive function.

**Figure 1.**
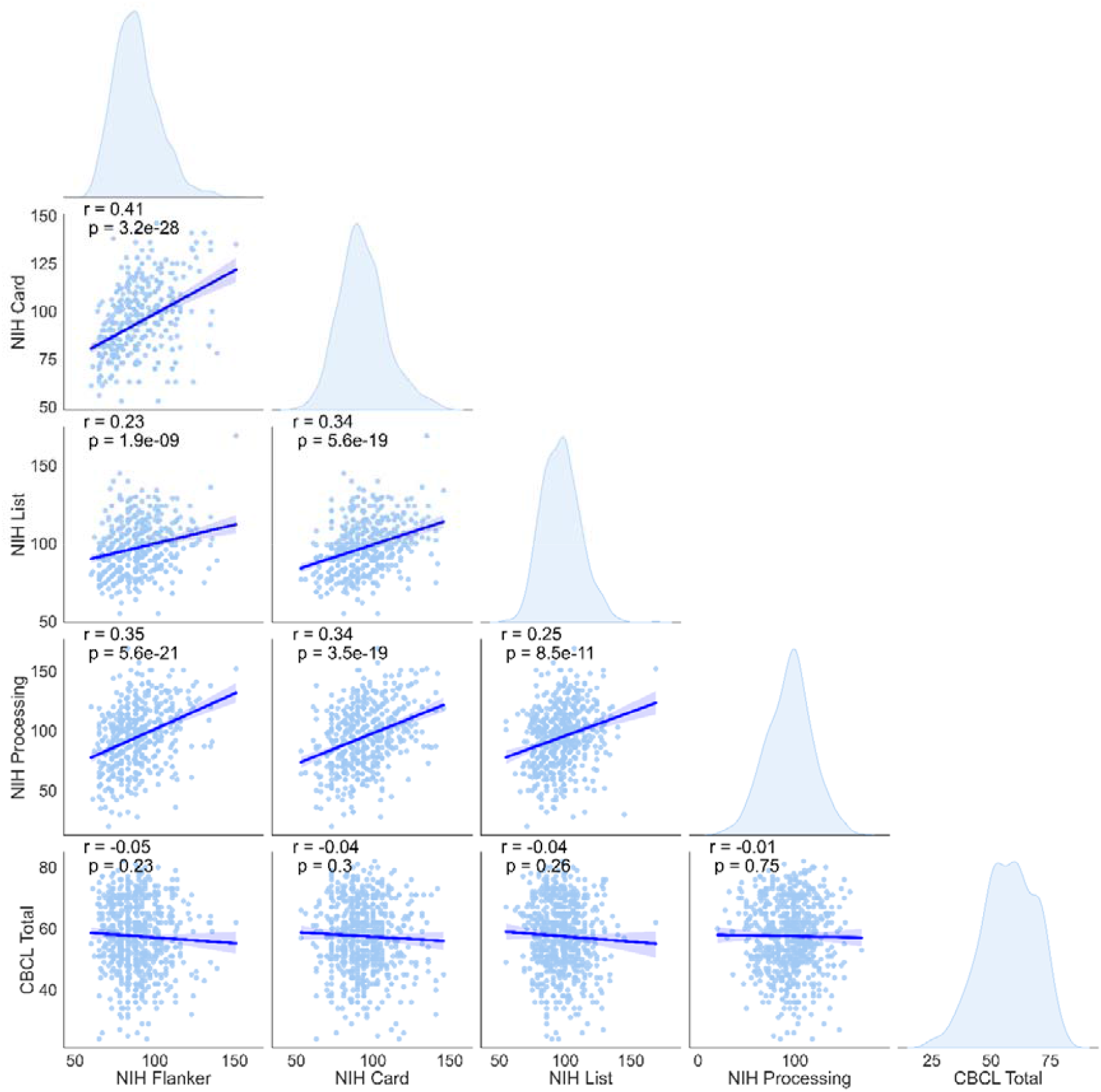
Scatterplot representation of the pair-wise correlation between psychopathology and cognitive characteristics (NIH toolbox subscales) scores. Histograms of the distribution of each variable with smooth curves obtained using a kernel density estimate function. *Abbreviations*: NIH Toolbox = NIH-TB; NIH List=NIH TB List Sorting Working Memory Test; NIH Card= NIH TB Cognition Domain Dimensional Change Card Sort Test; NIH Flanker = NIH TB Flanker Inhibitory Control and Attention Test; NIH Processing = NIH-TB Pattern Comparison Processing Speed Test; CBCL total = total t-score of Child Behavior Checklist.

**Figure 2.**
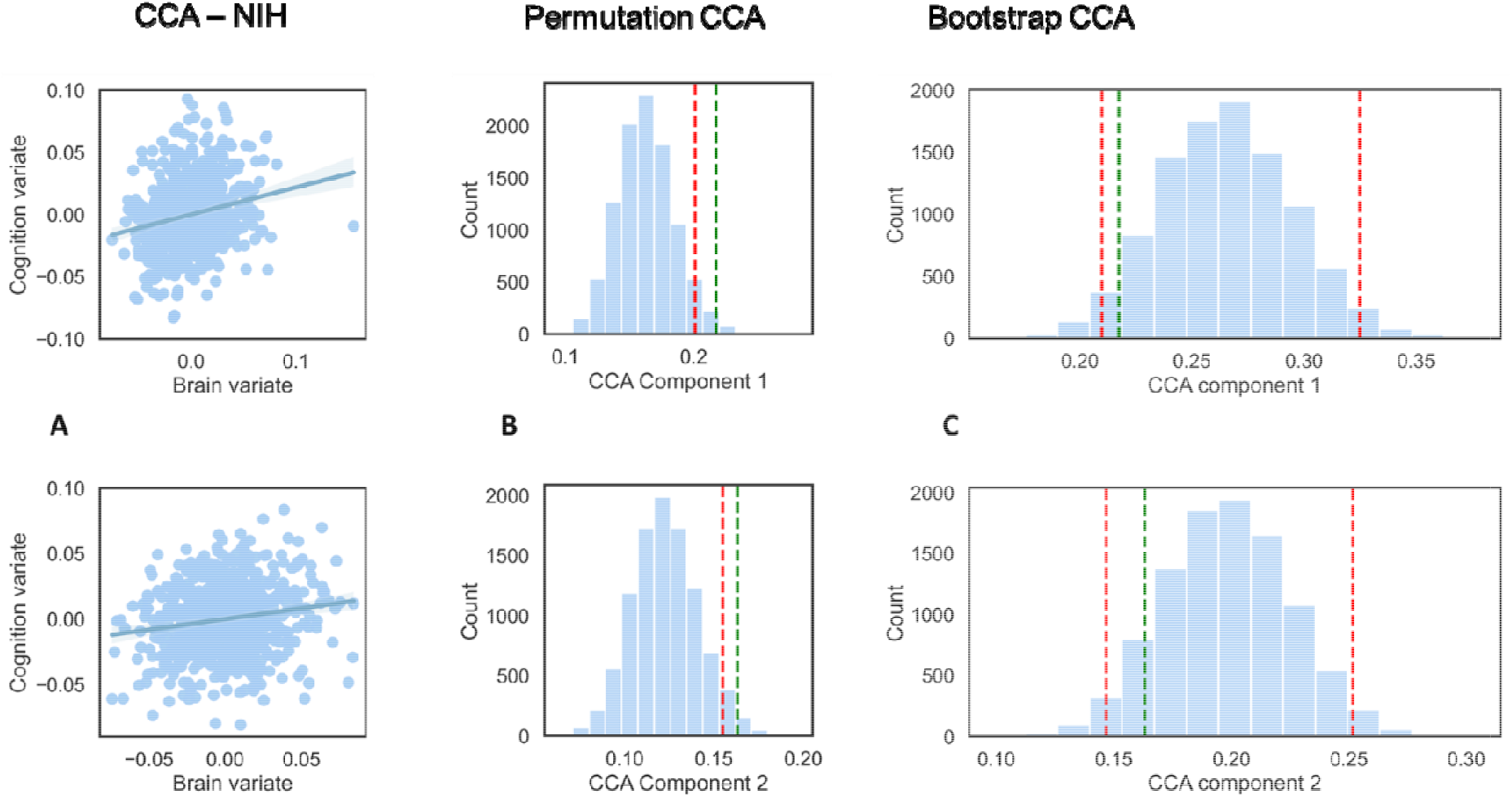
Results of the CCA analyses. A.1-2. Canonical correlation plots between the cerebellar and clinical variates. B.1 - B.2 Significance testing of the CCA. Distribution of CCA coefficients for component 1 (B.1) and component 2 (B.2) obtained by performing 10’000 permutations. Red line represents a significance threshold set for an alpha level of 0.05. C.1-C.2. Stability testing of the CCA. Distribution of canonical correlation coefficients between cerebellar and clinical variates by bootstrapping procedure with 10 ‘000 tests at an alpha level of 0.05 for component 1 (C.1) and component 2 (C.2). Lower and upper bound corresponding to +/-1.96 SD in red dotted line

### Cerebellar correlates of cognitive function

We then turned to the brain to examine whether we could uncover latent neural and behavioral dimensions to our data with a multivariate CCA approach (including age, sex, scan location, total intracranial volume, cognition (NIH-TB subscales). Such an approach allowed us to identify two significant correlation between the first cognitive canonical variate (the subscales of the NIH toolbox Cognition domain) and the first brain canonical variate (regional cerebellar gray matter volume and intracranial volume) at r=.22, as well as between the second clinical canonical variate (the subscales of the NIH toolbox Cognition domain) and the second brain canonical variate (regional cerebellar gray matter volume and intracranial volume) at r=.16. To assess significance, we conducted permutation testing with 10’000 tests at 95% that revealed that the correlation of both components was significant (statistical threshold for component 1: r = .20, and for component 2: r = .15) (Figure 2).

Next we performed a bootstrapping analysis to examine the stability of our multivariate CCA model (Sauerbrei and Royston 2007). Our results showed that our CCA results are stable and non-zero within the [5:95] confidence interval of the results generated by bootstrapping analysis.

Regarding the first component, cognitive flexibility (indexed by the NIH card subscale), processing speed (indexed by NIH processing scale) and working memory (indexed by the NIH list subscale) loaded the most on the clinical canonical variate at 0.89 (large effect), 0.65 and 0.52 (moderate effects), respectively. Gray matter volume in the Crus II and Lobule X loaded with a moderate effect size on the brain canonical variate at 0.57 and 0.59, respectively (Figure 3). Regarding the second component, working memory (indexed by the NIH list subscale) and cognitive control (indexed by NIH Flanker subscale) loaded with a moderate effect on the cognitive canonical variate at 0.51 and -0.56 respectively. Gray matter volume in the Crus I and Lobule VI loaded also moderately on the brain canonical variate at 0.49 each. Overall, these results

**Figure 3.**
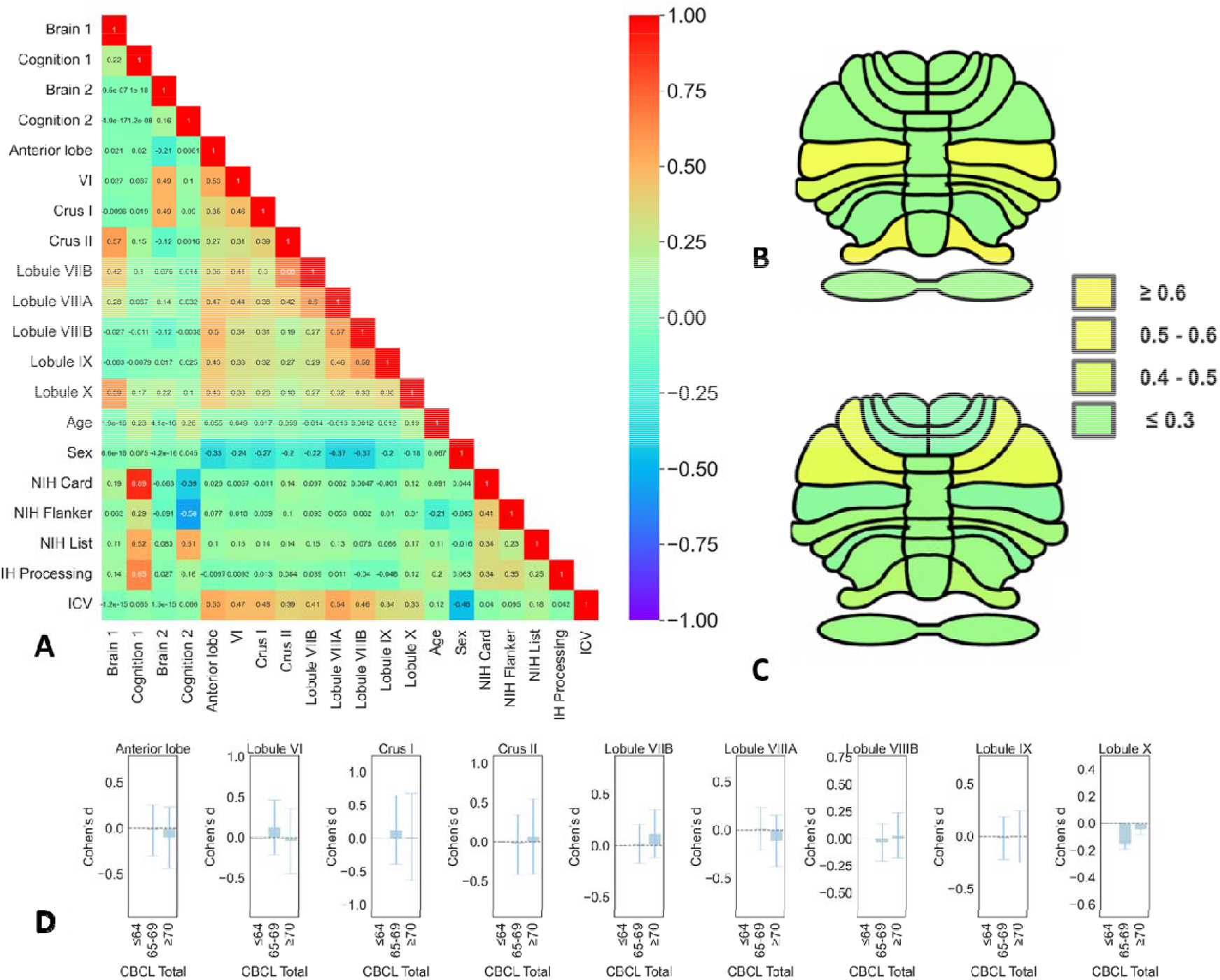
Relationship of cerebellar structure with cognition and psychopathology. A-C. Canonical correlation analysis: loading of cognitive and anatomical variates. D. Effect sizes as standardized mean difference in groups stratified by psychopathology severity quantified by CBCL t-score. *Annotations*: Brain 1 = first anatomical component; Brain 2 = second anatomical component; Cognition 1 = first cognitive component; Cognition 2 = second cognitive component; NIH Toolbox = NIH-TB; NIH List=NIH TB List Sorting Working Memory Test; NIH Card= NIH TB Cognition Domain Dimensional Change Card Sort Test; NIH Flanker = NIH TB Flanker Inhibitory Control and Attention Test; NIH Processing = NIH-TB Pattern Comparison Processing Speed Test; ICV=Intracranial Volume; CBCL= Child Behavior Checklist total t-score.

We then asked whether there the differences in cerebellar volumes observed with cognitive function could be explained by psychopathology severity. To address this, we stratified the total CBCL t-score in the established normative cut-offs of the CBCL with a t-score of ≤ 64 for non-clinical symptoms, a t-score between 65 and 69 for borderline individuals with risk for problem behaviors, and a t-score ≥ 70 for clinical symptoms (Achenbach and Rescorla 2001; Carta et al. 2020). We then computed the effect size (Cohen’s d) of differences in cerebellar volume using standardized mean differences accounting for age, sex, scan location and intracranial volume. We found no significant differences in cerebellar volumes across these categories (Figure 3D) showing a lack of psychopathology effects in cerebellar anatomy. Furthermore, we included the CBCL total t-scores and the NIH subscales in the same CCA model with permutation and bootstrapping. This analysis did not impact our main cognitive-cerebellum CCA results remaining both statistically significant and stable (Supplementary material, Figure S3). We asked then whether specific dimensions of psychopathology could drive cerebellar changes and performed CCA analyses with the subscales of the CBCL. These results were not statistically significant and did not survive bootstrapping (Supplementary material, Figure S4). In sum, our results indicated that cognitive function but not psychopathology severity drove the observed cerebellar changes.

## Discussion

Cognitive neuroscience is only beginning to unravel the role of the cerebellum in higher “supratentorial” cognitive functions. Despite historically being framed as a “motor control” brain region, extensive human neuroimaging and lesion evidence has suggested a cerebellar role in cognitive function. Furthermore, previous evidence uncovered a correspondence of cerebellar anatomy with *general* cognition and psychopathology. Therefore, we set out to examine for the first time the cerebellar topography in connection to specific components of cognition. Our multivariate analyses (CCA) outlined how different components of cognitive function map onto cerebellar morphometry independently of psychopathology severity in support of the cerebellar cognitive and affective syndrome (Schmahmann and Sherman 1998; Jacobi et al. 2021). In particular, we showed only partly shared cerebellar maps of cognitive function (Figure 3, A-C): a first map encompassing cognitive flexibility (large effect size) and speed of processing (moderate effect size) associated with regional gray matter volume in Crus II and Lobule X and a second map including the Crus I and Lobule VI associated with cognitive control (moderate effect size). Working memory associations were similarly present in both these maps (Crus II, Lobule X, Crus I and Lobule VI) with similar moderate effect sizes. These results account for psychopathology severity and other confounds and suggest that such correspondence between cerebellar anatomy may go across transdiagnostic boundaries. Crucially, permutation testing and bootstrapping analyses showed that these relationships are significant and our CCA model is robust and stable (Sauerbrei and Royston 2007).

Our findings highlight an association of the cerebellar Crus II and Lobule X structure with cognitive flexibility with a large effect size. Importantly, we show that cerebellar gray matter volume is linked to cognitive flexibility abilities in a dimensional across-diagnostic categories manner. While theoretical accounts have posited a role for the cerebellum in the flexible coordination of cognitive processes (Cognitive Dysmetria theory), strikingly, no large-scaled evidence for such contribution existed prior to our work. Animal research evidence indicated that hemi-cerebellectomized animals are unable to flexibly switch to a new set of rules, despite having intact motor responses (De Bartolo et al. 2009; Dickson et al. 2017). A few small-sample human studies (Ben-Soussan et al. 2015; Kansal et al. 2017; Badaly et al. 2022; Kühn et al. 2012; Paradiso et al. 1997; Parker et al. 2008; Koppelmans et al. 2017; Bauer et al. 2009; Bernard and Seidler 2013) have underlined a cerebellar role in mediating cognitive flexibility. The cerebellar correspondence of cognitive flexibility fits well with evidence from cerebellar lesions (Stoodley and Schmahmann 2010; Argyropoulos et al. 2020; Stoodley et al. 2016) and theoretical accounts of cognitive dysmetria theory (Andreasen, Paradiso, and O’Leary 1998). According to this theory, the cerebellum plays a key role in coordinating different cognitive and affective processes, quite similarly to its role in motor coordination. Impairments in cognitive flexibility seem to be prevalent in a variety of psychiatric disorders across the lifespan (O’Donnell et al. 2017; Dotson et al. 2020; Goodall et al. 2018; Geurts, Corbett, and Solomon 2009; Verdejo-Garcia et al. 2015) and they represent a potentially important pharmacological (Doss et al. 2021), psychotherapeutic (Johnco, Wuthrich, and Rapee 2014), or neuromodulation (de Boer et al. 2021) target.

Our results point to two cognitive-anatomical maps both related to working memory in agreement with previous findings (Hayter, Langdon, and Ramnani 2007; Guell, Gabrieli, and Schmahmann 2018; Marvel and Desmond 2016; Ashida et al. 2019; Begue et al. 2022). Furthermore, we show a first component characterizing the positive association of cognitive flexibility and speed of processing with regional gray matter volume in Crus II and Lobule X and a second, separate, component that captures the relationship of Crus I and Lobule VI with cognitive control (negative association). Speed of processing (the ability to quickly process information), working memory (the ability to hold and manipulate information during short periods of time) and cognitive flexibility (the ability to switch rapidly between mental states and tasks) are interconnected cognitive capacities that are important for flexible behavior. On the other hand, cognitive stability - the ability to maintain stable cognitive representations is also important for consistent and reliable behavior, and requires less task-switching and more working memory capacity. Previous literature has focused on supratentorial prefronto-striatal networks of working memory, cognitive flexibility and stability (Cools and D’Esposito 2010; Cools 2016; Westbrook and Braver 2016). In this circuitry, cognitive flexibility and cognitive stability have been shown to rely on dopaminergic signaling in the striatum, as demonstrated in human PET neuroimaging (Klostermann et al. 2012; Rieckmann et al. 2011; Landau et al. 2009) and task-related studies (Berry et al. 2016; Samanez-Larkin et al. 2013), and in the dorsolateral prefrontal cortices (Borwick et al. 2020). In line with these findings, Braver and Westbrook (Westbrook and Braver 2016) attributed a key role to dopaminergic neurons in the flexible coordination of cognitive processes for goal-directed behavior. In other words, during goal-directed behavior, phasic dopamine release promotes updating of these representations and therefore cognitive flexibility, whereas tonic dopamine release maintains the stability of cognitive representations. In light of our results, we speculatively propose that, similarly, dopaminergic signaling may underlie cerebellar participation in cognitive flexibility or stability either through direct local dopaminergic signaling in the cerebellum or *via* indirect (e.g., distant) cerebellar prefrontal closed loops as part of “the rich club” (Watson et al. 2014). Indeed, high levels of dopamine have been found in postmortem cerebellum of humans, rats, and monkeys reviewed in (Flace et al. 2021). Further research is needed to examine whether and how dopaminergic signaling underlies cerebellar correlates of cognitive function.

Our study has several strengths. First, we overcome previous shortcomings of case-control studies by endorsing a dimensional approach that captures phenotypic gradients in a large cohort. To our knowledge, this is the largest study to date to ever examine cerebellar contributions to distinct cognitive components including cognitive flexibility. Second, we employ a validated pipeline (CERES) with careful and stringent quality control to ensure optimal preprocessing and avoid spurious results. In the current work, we have included only high-quality imaging data surviving a stringent visual quality check (e.g., only 662 of the images have passed the quality check of the initial n = 1452 subjects, Figure S1) using the same quality - control protocol employed previously (Laidi et al. 2022). Third, this study provides statistically significant (permutation testing) and stable (bootstrapping) data-driven results in the largest transdiagnostic sample to date to ascertain the significance of the results and the stability of the CCA model. Permutation testing allowed us to establish that the obtained results are statistically significant. However, obtaining significant results does not exclude the possibility of having a random sampling error (e.g., the sample does not represent the general population). Such a possibility can be ruled out by bootstrapping: the results of the original cohort can be compared with bootstrapped samples, allowing us to examine the robustness of the model. Here, our results survived bootstrapping and we can confidently state that our results are not due to a random sampling error. Regarding limitations, our cross-sectional design and correlational analyses prevent inferences on the causal nature of the observed changes in the cerebellum. Furthermore, the findings may not generalize to older populations considering that in this cohort only young developing individuals (mean age of 10.5 years) were included. Future follow-up studies should examine how these relationships are expressed in adult cohorts.

Our work links cerebellar morphometry to distinct components of cognitive function including cognitive flexibility. These functions are observed to be altered in psychiatric disorders such as schizophrenia, depression, autism, and obsessive-compulsive disorders (Dajani and Uddin 2015), all of which also are shown to have cerebellar aberrations (Phillips et al. 2015). Given the recent advance in cerebellar non-invasive brain stimulation and its association with neuroimaging (Brady et al. 2019; Yao et al. 2022), our work opens the perspective of cerebellar targeting across different psychiatric diagnoses for cognitive improvement. Overall, our results elucidate for the first time the cerebellar anatomical circuitry supporting inter-individual differences in cognitive function and highlight a prominent role for the human cerebellum in distinct aspects of cognition for flexible adaptive behavior.

## Data Availability

data are available on request from the authors.

## Supplementary material

### Supplementary Material 1. Linear univariate analyses

We also examined post-hoc linear univariate relationships of cerebellar regional gray matter to cognitive abilities measured by NIH toolbox subscales controlling for age, ICV, and psychopathology severity (indexed by CBCL total scores) as continuous variables and sex, scan location as categorical variables (Figure S2). We found a main positive effect of NIH List subscale indexing working memory in gray matter volumes of the anterior lobe (p=0. 015), Lobules VI (p = 0.0008), Crus I (p = 0.0008), Crus II (p =0.001), VIIB (p =0.0008), VIIIA (p=0.003) and X (p = 0.0008). Further, we found a main positive effect of NIH Card indexing cognitive flexibility in Crus II (p =.001), VIIB (p=.015), VIIIA (p=0.03) and Lobule X (p=.004). Cognitive control (indexed by NIH Flanker’s subscore) correlated with gray matter volume in the anterior lobe gray matter volume (p=0.04) and Crus II (p=0.03) whereas processing speed (indexed by NIH processing subscore) positively correlated to gray matter volume in Crus II (p=0.04) and Lobule X (p= 0.04) (all p-values are FDR corrected).

**Figure S1.**
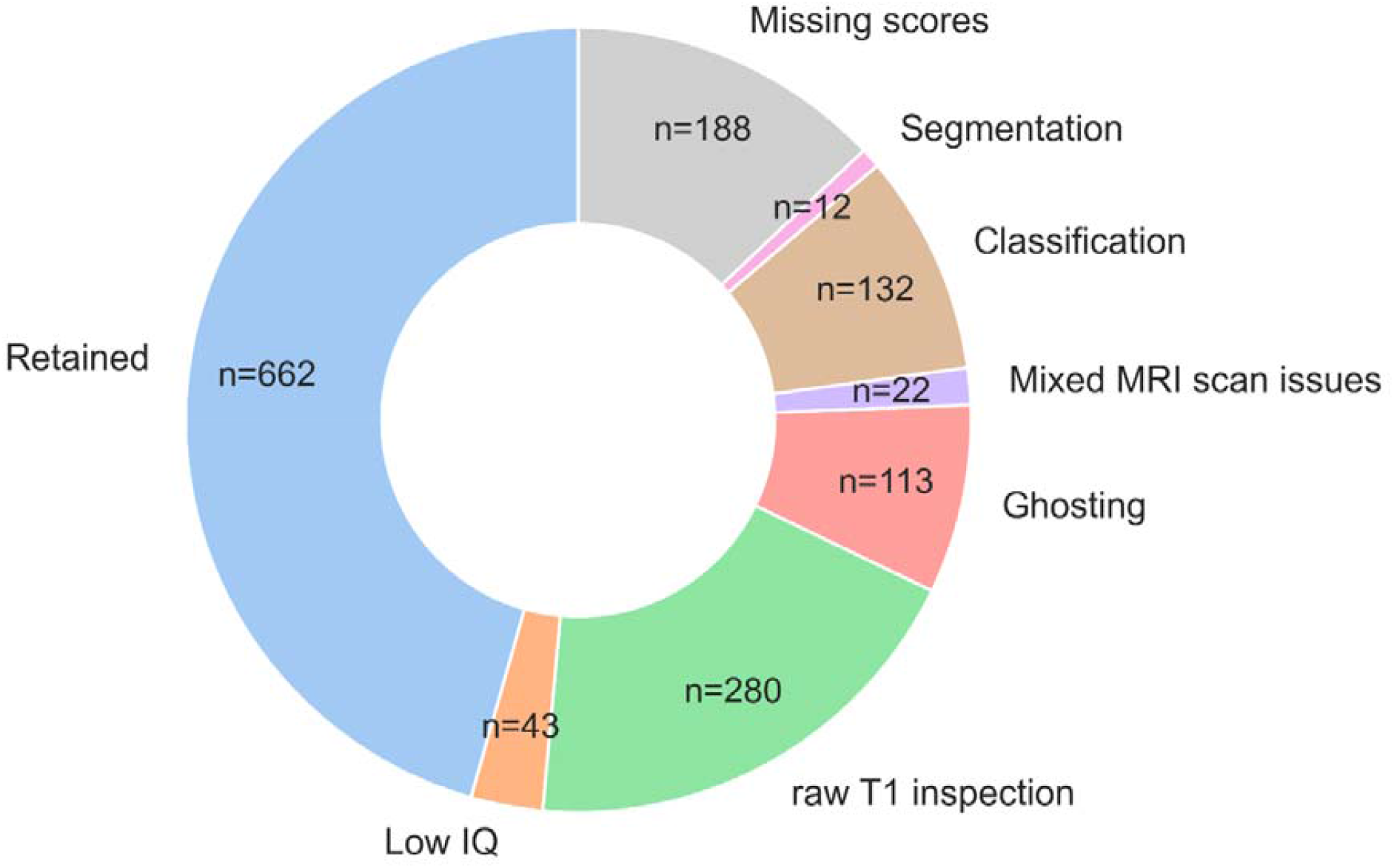
Repartition of excluded individuals from the final analysis and the respective percentages.

**Figure S2.**
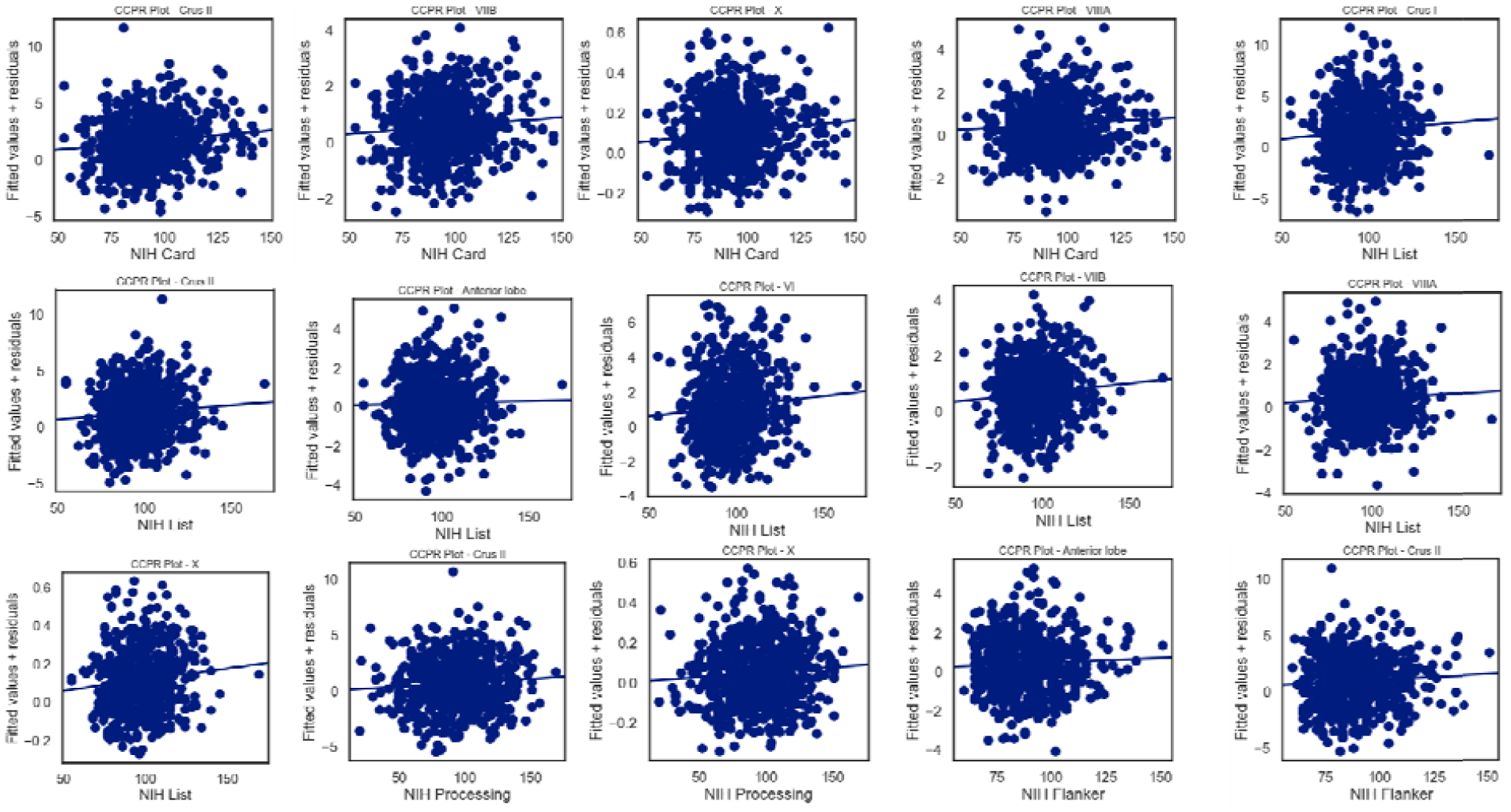
Summary of univariate analysis statistically significant results (p<0.05 FDR corrected). Component and component-residual (CCPR) plots of NIH subscales with cerebellar regional gray matter volume

**Figure S3.**
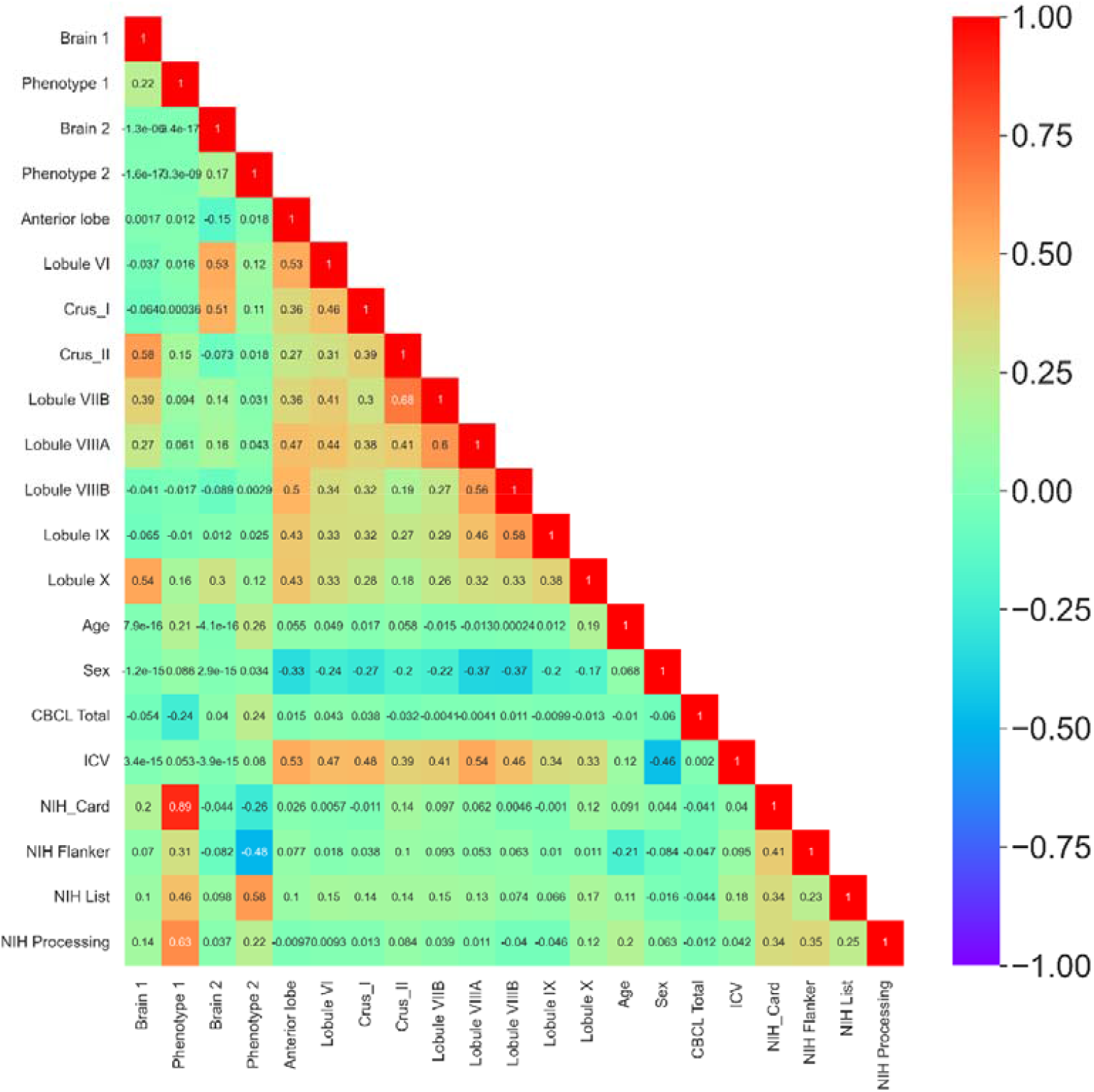
Correlation matrix of the CCA results of the relationship of cerebellar structure with cognition and psychopathology. *Annotations*: Brain 1 = first anatomical component; Brain 2 = second anatomical component; Cognition 1 = first cognitive component; Cognition 2 = second cognitive component; NIH Toolbox = NIH-TB; NIH List=NIH TB List Sorting Working Memory Test; NIH Card= NIH TB Cognition Domain Dimensional Change Card Sort Test; NIH Flanker = NIH TB Flanker Inhibitory Control and Attention Test; NIH Processing = NIH-TB Pattern Comparison Processing Speed Test; ICV=Intracranial Volume; CBCL total= Child Behavior Checklist total t-score.

**Figure S4.**
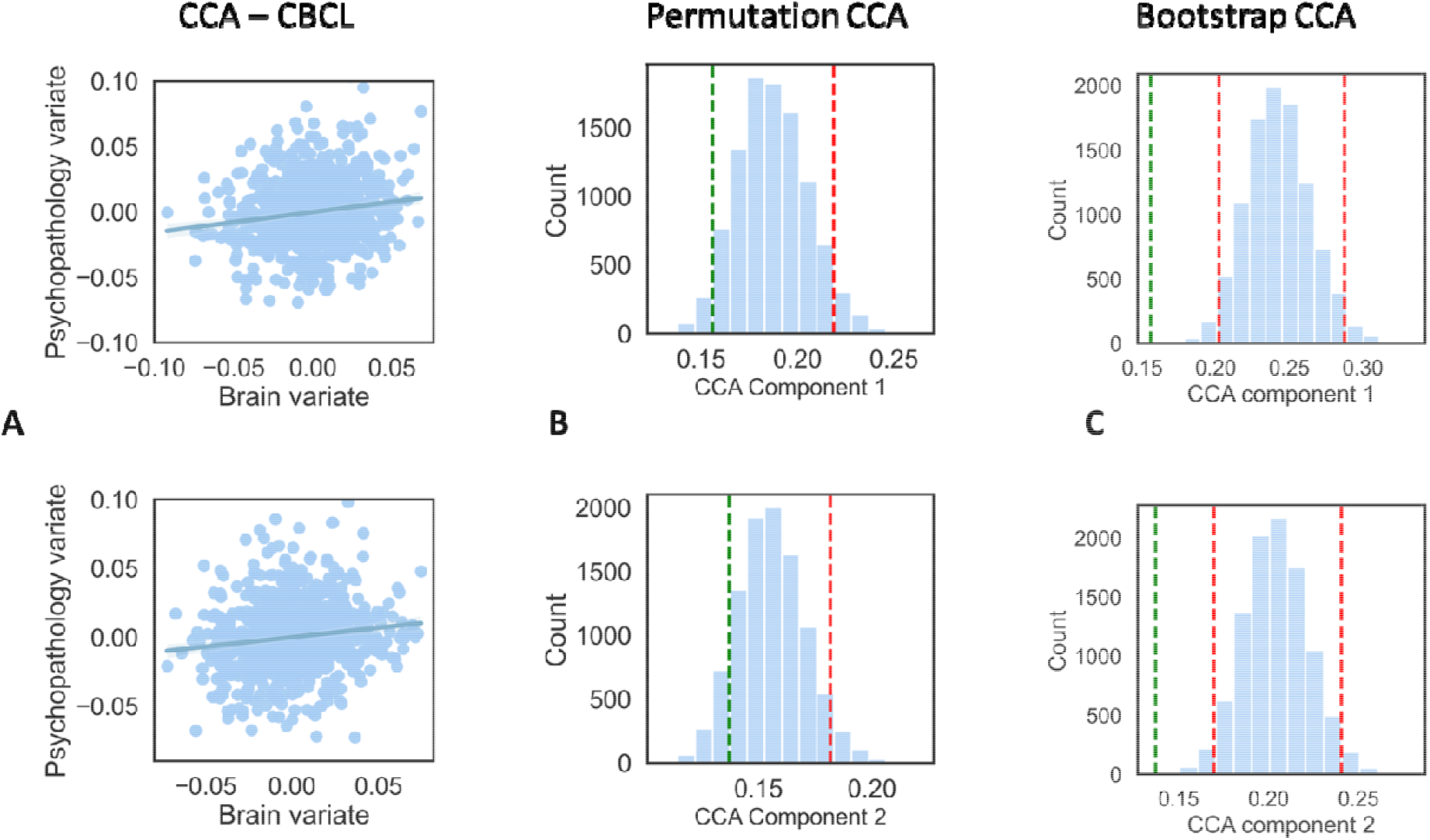
Results of the CCA with CBCL subscores. A. Canonical correlation plots between the cerebellar and clinical variates for the first component (top panel) and the second (bottom panel) B. Significance testing of the CCA. Distribution of CCA coefficients for component 1 (top) and component 2 (bottom) obtained by performing 10’000 permutations. Red line represents a significance threshold set for an alpha level of 0.05. C. Stability testing of the CCA. Distribution of canonical correlation coefficients between cerebellar and clinical variates by bootstrapping procedure with 10 ‘000 tests at an alpha level of 0.05 for component 1 (top) and component 2 (bottom). Lower and upper bound corresponding to +/-1.96 SD in red dotted line

## References

Achenbach, Thomas M., and Leslie A. Rescorla. 2001. Manual for the ASEBA School-Age Forms & Profiles: An Integrated System of Multi-Informant Assessment. ASEBA.

Alexander, Lindsay M., Jasmine Escalera, Lei Ai, Charissa Andreotti, Karina Febre, Alexander Mangone, Natan Vega-Potler, et al. 2017. “An Open Resource for Transdiagnostic Research in Pediatric Mental Health and Learning Disorders.” Scientific Data 4 (1): 170181. https://doi.org/10.1038/sdata.2017.181.

Andreasen, N. C., S. Paradiso, and D. S. O’Leary. 1998. “‘Cognitive Dysmetria’ as an Integrative Theory of Schizophrenia: A Dysfunction in Cortical-Subcortical-Cerebellar Circuitry?” Schizophrenia Bulletin 24 (2): 203–18. https://doi.org/10.1093/oxfordjournals.schbul.a033321.

Argyropoulos, G. P. D., K. van Dun, M. Adamaszek, M. Leggio, M. Manto, M. Masciullo, M. Molinari, et al. 2020. “The Cerebellar Cognitive Affective/Schmahmann Syndrome: A Task Force Paper.” Cerebellum 19 (1): 102–25. https://doi.org/10.1007/s12311-019-01068-8.

Ashida, Reiko, Nadia L. Cerminara, Richard J. Edwards, Richard Apps, and Jonathan C. W. Brooks. 2019. “Sensorimotor, Language, and Working Memory Representation within the Human Cerebellum.” Human Brain Mapping 40 (16): 4732–47. https://doi.org/10.1002/hbm.24733.

Badaly, Daryaneh, Sue R. Beers, Rafael Ceschin, Vincent K. Lee, Shahida Sulaiman, Alexandria Zahner, Julia Wallace, et al. 2022. “Cerebellar and Prefrontal Structures Associated With Executive Functioning in Pediatric Patients With Congenital Heart Defects.” Frontiers in Neurology 13. https://www.frontiersin.org/articles/10.3389/fneur.2022.827780.

Bauer, Patrick M., Jamie L. Hanson, Ronald K. Pierson, Richard J. Davidson, and Seth D. Pollak. 2009. “Cerebellar Volume and Cognitive Functioning in Children Who Experienced Early Deprivation.” Biological Psychiatry 66 (12): 1100–1106. https://doi.org/10.1016/j.biopsych.2009.06.014.

Begue, Indrit, Janis Brakowski, Erich Seifritz, Alain Dagher, Philippe N. Tobler, Matthias Kirschner, and Stefan Kaiser. 2022. “Cerebellar and Cortico-Striatal-Midbrain Contributions to Reward-Cognition Processes and Apathy within the Psychosis Continuum.” SCHIZOPHRENIA RESEARCH 246 (August): 85–94. https://doi.org/10.1016/j.schres.2022.06.010.

Ben-Soussan, Tal D., Aviva Berkovich-Ohana, Claudia Piervincenzi, Joseph Glicksohn, and Filippo Carducci. 2015. “Embodied Cognitive Flexibility and Neuroplasticity Following Quadrato Motor Training.” Frontiers in Psychology 6. https://www.frontiersin.org/articles/10.3389/fpsyg.2015.01021.

Bernard, Jessica A., and Rachael D. Seidler. 2013. “Relationships between Regional Cerebellar Volume and Sensorimotor and Cognitive Function in Young and Older Adults.” Cerebellum (London, England) 12 (5): 721–37. https://doi.org/10.1007/s12311-013-0481-z.

Berry, Anne S., Vyoma D. Shah, Suzanne L. Baker, Jacob W. Vogel, James P. O’Neil, Mustafa Janabi, Henry D. Schwimmer, Shawn M. Marks, and William J. Jagust. 2016. “Aging Affects Dopaminergic Neural Mechanisms of Cognitive Flexibility.” Journal of Neuroscience 36 (50): 12559–69. https://doi.org/10.1523/JNEUROSCI.0626-16.2016.

Bilenko, Natalia Y., and Jack L. Gallant. 2016. “Pyrcca: Regularized Kernel Canonical Correlation Analysis in Python and Its Applications to Neuroimaging.” Frontiers in Neuroinformatics 10. https://www.frontiersin.org/articles/10.3389/fninf.2016.00049.

Boer, Nina S. de, Renée S. Schluter, Joost G. Daams, Ysbrand D. van der Werf, Anna E. Goudriaan, and Ruth J. van Holst. 2021. “The Effect of Non-Invasive Brain Stimulation on Executive Functioning in Healthy Controls: A Systematic Review and Meta-Analysis.” Neuroscience & Biobehavioral Reviews 125 (June): 122–47. https://doi.org/10.1016/j.neubiorev.2021.01.013.

Borwick, Ciara, Reece Lal, Lee Wei Lim, Charlotte J. Stagg, and Luca Aquili. 2020. “Dopamine Depletion Effects on Cognitive Flexibility as Modulated by TDCS of the DlPFC.” Brain Stimulation 13 (1): 105–8. https://doi.org/10.1016/j.brs.2019.08.016.

Brady, Roscoe O., Irene Gonsalvez, Ivy Lee, Dost Öngür, Larry J. Seidman, Jeremy D. Schmahmann, Shaun M. Eack, Matcheri S. Keshavan, Alvaro Pascual-Leone, and Mark A. Halko. 2019. “Cerebellar-Prefrontal Network Connectivity and Negative Symptoms in Schizophrenia.” The American Journal of Psychiatry 176 (7): 512–20. https://doi.org/10.1176/appi.ajp.2018.18040429.

Buckner, Randy L., Fenna M. Krienen, Angela Castellanos, Julio C. Diaz, and B. T. Thomas Yeo. 2011. “The Organization of the Human Cerebellum Estimated by Intrinsic Functional Connectivity.” Journal of Neurophysiology 106 (5): 2322–45. https://doi.org/10.1152/jn.00339.2011.

Carass, Aaron, Jennifer L. Cuzzocreo, Shuo Han, Carlos R. Hernandez-Castillo, Paul E. Rasser, Melanie Ganz, Vincent Beliveau, et al. 2018. “Comparing Fully Automated State-of-the-Art Cerebellum Parcellation from Magnetic Resonance Images.” NeuroImage 183 (December): 150–72. https://doi.org/10.1016/j.neuroimage.2018.08.003.

Carpenter, J., and J. Bithell. 2000. “Bootstrap Confidence Intervals: When, Which, What? A Practical Guide for Medical Statisticians.” Statistics in Medicine 19 (9): 1141–64. https://doi.org/10.1002/(sici)1097-0258(20000515)19:9<1141::aid-sim479>3.0.co;2-f.

Carta, Alessandra, Elisa Fucà, Silvia Guerrera, Eleonora Napoli, Giovanni Valeri, and Stefano Vicari. 2020. “Characterization of Clinical Manifestations in the Co-Occurring Phenotype of Attention Deficit/Hyperactivity Disorder and Autism Spectrum Disorder.” Frontiers in Psychology 11 (May): 861. https://doi.org/10.3389/fpsyg.2020.00861.

Chavez-Baldini, UnYoung, Dorien H. Nieman, Amos Keestra, Anja Lok, Roel J. T. Mocking, Pelle de Koning, Valeria V. Krzhizhanovskaya, et al. 2021. “The Relationship between Cognitive Functioning and Psychopathology in Patients with Psychiatric Disorders: A Transdiagnostic Network Analysis.” Psychological Medicine 53 (2): 1–10. https://doi.org/10.1017/S0033291721001781.

Cools, Roshan. 2016. “The Costs and Benefits of Brain Dopamine for Cognitive Control.” Wiley Interdisciplinary Reviews: Cognitive Science 7 (5): 317–29.

Cools, Roshan, and Mark D’Esposito. 2010. “Dopaminergic Modulation of Flexible Cognitive Control in Humans.” Dopamine Handbook, January. https://doi.org/10.1093/acprof:oso/9780195373035.003.0017.

Cuthbert, Bruce N. 2014. “The RDoC Framework: Facilitating Transition from ICD/DSM to Dimensional Approaches That Integrate Neuroscience and Psychopathology.” World Psychiatry 13 (1): 28–35. https://doi.org/10.1002/wps.20087.

Dajani, Dina R., and Lucina Q. Uddin. 2015. “Demystifying Cognitive Flexibility: Implications for Clinical and Developmental Neuroscience.” Trends in Neurosciences 38 (9): 571–78. https://doi.org/10.1016/j.tins.2015.07.003.

De Bartolo, P., L. Mandolesi, F. Federico, F. Foti, D. Cutuli, F. Gelfo, and L. Petrosini. 2009. “Cerebellar Involvement in Cognitive Flexibility.” Neurobiology of Learning and Memory 92 (3): 310–17. https://doi.org/10.1016/j.nlm.2009.03.008.

Dickson, P. E., J. Cairns, D. Goldowitz, and G. Mittleman. 2017. “Cerebellar Contribution to Higher and Lower Order Rule Learning and Cognitive Flexibility in Mice.” Neuroscience 345 (March): 99–109. https://doi.org/10.1016/j.neuroscience.2016.03.040.

Doss, Manoj K., Michal Považan, Monica D. Rosenberg, Nathan D. Sepeda, Alan K. Davis, Patrick H. Finan, Gwenn S. Smith, et al. 2021. “Psilocybin Therapy Increases Cognitive and Neural Flexibility in Patients with Major Depressive Disorder.” Translational Psychiatry 11 (1): 1–10. https://doi.org/10.1038/s41398-021-01706-y.

Dotson, Vonetta M., Shawn M. McClintock, Paul Verhaeghen, Joseph U. Kim, Amanda A. Draheim, Sarah M. Syzmkowicz, Andrew M. Gradone, Hannah R. Bogoian, and Liselotte De Wit. 2020. “Depression and Cognitive Control across the Lifespan: A Systematic Review and Meta-Analysis.” Neuropsychology Review 30 (4): 461–76. https://doi.org/10.1007/s11065-020-09436-6.

Ericson, Thomas, and Victor Zinoviev. 2001. “Chapter 4 - Permutation Codes.” In North-Holland Mathematical Library, edited by Thomas Ericson and Victor Zinoviev, 63:107–27. Codes on Euclidean Spheres. Elsevier. https://doi.org/10.1016/S0924-6509(01)80049-0.

Flace, P., P. Livrea, G.A. Basile, D. Galletta, A. Bizzoca, G. Gennarini, S. Bertino, et al. 2021. “The Cerebellar Dopaminergic System.” Frontiers in Systems Neuroscience 15 ((Flace P.) Medical School, University of Bari ‘Aldo Moro’, Bari, Italy). https://doi.org/10.3389/fnsys.2021.650614.

Geurts, Hilde M., Blythe Corbett, and Marjorie Solomon. 2009. “The Paradox of Cognitive Flexibility in Autism.” Trends in Cognitive Sciences 13 (2): 74–82. https://doi.org/10.1016/j.tics.2008.11.006.

Goodall, Joanne, Caroline Fisher, Sarah Hetrick, Lisa Phillips, Emma M. Parrish, and Kelly Allott. 2018. “Neurocognitive Functioning in Depressed Young People: A Systematic Review and Meta-Analysis.” Neuropsychology Review 28 (2): 216–31. https://doi.org/10.1007/s11065-018-9373-9.

Guell, Xavier, John D. E. Gabrieli, and Jeremy D. Schmahmann. 2018. “Triple Representation of Language, Working Memory, Social and Emotion Processing in the Cerebellum: Convergent Evidence from Task and Seed-Based Resting-State FMRI Analyses in a Single Large Cohort.” NeuroImage 172 (May): 437–49. https://doi.org/10.1016/j.neuroimage.2018.01.082.

Guell, Xavier, Jeremy D Schmahmann, John DE Gabrieli, and Satrajit S Ghosh. 2018. “Functional Gradients of the Cerebellum.” Edited by Andreea Bostan and Richard B Ivry. ELife 7 (August): e36652. https://doi.org/10.7554/eLife.36652.

Hardoon, David R., Janaina Mourão-Miranda, Michael Brammer, and John Shawe-Taylor. 2007. “Unsupervised Analysis of FMRI Data Using Kernel Canonical Correlation.” NeuroImage 37 (4): 1250–59. https://doi.org/10.1016/j.neuroimage.2007.06.017.

Hayter, A. L., D. W. Langdon, and N. Ramnani. 2007. “Cerebellar Contributions to Working Memory.” NeuroImage 36 (3): 943–54. https://doi.org/10.1016/j.neuroimage.2007.03.011.

Jacobi, Heike, Jennifer Faber, Dagmar Timmann, and Thomas Klockgether. 2021. “Update Cerebellum and Cognition.” Journal of Neurology 268 (10): 3921–25. https://doi.org/10.1007/s00415-021-10486-w.

Johnco, C., V. M. Wuthrich, and R. M. Rapee. 2014. “The Influence of Cognitive Flexibility on Treatment Outcome and Cognitive Restructuring Skill Acquisition during Cognitive Behavioural Treatment for Anxiety and Depression in Older Adults: Results of a Pilot Study.” Behaviour Research and Therapy 57 (June): 55–64. https://doi.org/10.1016/j.brat.2014.04.005.

Kansal, Kalyani, Zhen Yang, Ann M. Fishman, Haris I. Sair, Sarah H. Ying, Bruno M. Jedynak, Jerry L. Prince, and Chiadi U. Onyike. 2017. “Structural Cerebellar Correlates of Cognitive and Motor Dysfunctions in Cerebellar Degeneration.” Brain 140 (3): 707–20. https://doi.org/10.1093/brain/aww327.

King, Maedbh, Carlos R. Hernandez-Castillo, Russell A. Poldrack, Richard B. Ivry, and Jörn Diedrichsen. 2019. “Functional Boundaries in the Human Cerebellum Revealed by a Multi-Domain Task Battery.” Nature Neuroscience 22 (8): 1371–78. https://doi.org/10.1038/s41593-019-0436-x.

Klostermann, Ellen C., Meredith N. Braskie, Susan M. Landau, James P. O’Neil, and William J. Jagust. 2012. “Dopamine and Frontostriatal Networks in Cognitive Aging.” Neurobiology of Aging 33 (3): 623.e15-623.e24. https://doi.org/10.1016/j.neurobiolaging.2011.03.002.

Koppelmans, Vincent, Yoo Young Hoogendam, Sarah Hirsiger, Susan Mérillat, Lutz Jäncke, and Rachael D. Seidler. 2017. “Regional Cerebellar Volumetric Correlates of Manual Motor and Cognitive Function.” Brain Structure & Function 222 (4): 1929–44. https://doi.org/10.1007/s00429-016-1317-7.

Kühn, Simone, Alexander Romanowski, Florian Schubert, and Jürgen Gallinat. 2012. “Reduction of Cerebellar Grey Matter in Crus I and II in Schizophrenia.” Brain Structure & Function 217 (2): 523–29. https://doi.org/10.1007/s00429-011-0365-2.

Laidi, Charles, Dorothea L. Floris, Julian Tillmann, Yannis Elandaloussi, Mariam Zabihi, Tony Charman, Thomas Wolfers, et al. 2022. “Cerebellar Atypicalities in Autism?” Biological Psychiatry 92 (8): 674–82. https://doi.org/10.1016/j.biopsych.2022.05.020.

Landau, Susan M., Rayhan Lal, James P. O’Neil, Suzanne Baker, and William J. Jagust. 2009. “Striatal Dopamine and Working Memory.” Cerebral Cortex 19 (2): 445–54. https://doi.org/10.1093/cercor/bhn095.

Manjón, José V., Simon F. Eskildsen, Pierrick Coupé, José E. Romero, D. Louis Collins, and Montserrat Robles. 2014. “Nonlocal Intracranial Cavity Extraction.” International Journal of Biomedical Imaging 2014: 820205. https://doi.org/10.1155/2014/820205.

Marvel, Cherie L., and John E. Desmond. 2016. “Chapter 3 - The Cerebellum and Verbal Working Memory.” In The Linguistic Cerebellum, edited by Peter Mariën and Mario Manto, 51–62. San Diego: Academic Press. https://doi.org/10.1016/B978-0-12-801608-4.00003-7.

Mattoni, Matthew, Sylia Wilson, and Thomas M. Olino. 2021. “Identifying Profiles of Brain Structure and Associations with Current and Future Psychopathology in Youth.” Developmental Cognitive Neuroscience 51 (September): 101013. https://doi.org/10.1016/j.dcn.2021.101013.

Mazefsky, Carla A., Ranita Anderson, Caitlin M. Conner, and Nancy Minshew. 2011. “Child Behavior Checklist Scores for School-Aged Children with Autism: Preliminary Evidence of Patterns Suggesting the Need for Referral.” Journal of Psychopathology and Behavioral Assessment 33 (1): 31–37. https://doi.org/10.1007/s10862-010-9198-1.

Moberget, Torgeir, Dag Alnæs, Tobias Kaufmann, Nhat Trung Doan, Aldo Córdova-Palomera, Linn Bonaventure Norbom, Jaroslav Rokicki, Dennis van der Meer, Ole A. Andreassen, and Lars T. Westlye. 2019. “Cerebellar Gray Matter Volume Is Associated With Cognitive Function and Psychopathology in Adolescence.” Biological Psychiatry, Clinical Impact of Psychosis Risk Mechanisms, 86 (1): 65–75. https://doi.org/10.1016/j.biopsych.2019.01.019.

Moussa-Tooks, Alexandra B., Baxter P. Rogers, Anna S. Huang, Julia M. Sheffield, Stephan Heckers, and Neil D. Woodward. 2022. “Cerebellar Structure and Cognitive Ability in Psychosis.” Biological Psychiatry 92 (5): 385–95. https://doi.org/10.1016/j.biopsych.2022.03.013.

Na, Sabrina D., and Thomas G. Burns. 2016. “Wechsler Intelligence Scale for Children-V: Test Review.” Applied Neuropsychology. Child 5 (2): 156–60. https://doi.org/10.1080/21622965.2015.1015337.

O’Donnell, Lisa A., Patricia J. Deldin, Bethany Pester, Melvin G. McInnis, Scott A. Langenecker, and Kelly A. Ryan. 2017. “Cognitive Flexibility: A Trait of Bipolar Disorder That Worsens with Length of Illness.” Journal of Clinical and Experimental Neuropsychology 39 (10): 979–87. https://doi.org/10.1080/13803395.2017.1296935.

Paradiso, S., N. C. Andreasen, D. S. O’Leary, S. Arndt, and R. G. Robinson. 1997. “Cerebellar Size and Cognition: Correlations with IQ, Verbal Memory and Motor Dexterity.” Neuropsychiatry, Neuropsychology, and Behavioral Neurology 10 (1): 1–8.

Park, Min Tae M., Jon Pipitone, Lawrence H. Baer, Julie L. Winterburn, Yashvi Shah, Sofia Chavez, Mark M. Schira, et al. 2014. “Derivation of High-Resolution MRI Atlases of the Human Cerebellum at 3T and Segmentation Using Multiple Automatically Generated Templates.” NeuroImage 95 (July): 217–31. https://doi.org/10.1016/j.neuroimage.2014.03.037.

Parker, Jennifer, Ann Mitchell, Anastasia Kalpakidou, Muriel Walshe, Hee-Yeon Jung, Chiara Nosarti, Paramala Santosh, et al. 2008. “Cerebellar Growth and Behavioural & Neuropsychological Outcome in Preterm Adolescents.” Brain: A Journal of Neurology 131 (Pt 5): 1344–51. https://doi.org/10.1093/brain/awn062.

Patel, Yash, Nadine Parker, Giovanni A. Salum, Zdenka Pausova, and Tomáš Paus. 2022. “General Psychopathology, Cognition, and the Cerebral Cortex in 10-Year-Old Children: Insights From the Adolescent Brain Cognitive Development Study.” Frontiers in Human Neuroscience 15. https://www.frontiersin.org/articles/10.3389/fnhum.2021.781554.

Pedregosa, Fabian, Gaël Varoquaux, Alexandre Gramfort, Vincent Michel, Bertrand Thirion, Olivier Grisel, Mathieu Blondel, Peter Prettenhofer, Ron Weiss, and Vincent Dubourg. 2011. “Scikit-Learn: Machine Learning in Python.” The Journal of Machine Learning Research 12: 2825–30.

Phillips, Joseph R., Doaa H. Hewedi, Abeer M. Eissa, and Ahmed A. Moustafa. 2015. “The Cerebellum and Psychiatric Disorders.” Frontiers in Public Health 3 (May): 66. https://doi.org/10.3389/fpubh.2015.00066.

Rieckmann, Anna, Sari Karlsson, Håkan Fischer, and Lars Bäckman. 2011. “Caudate Dopamine D1 Receptor Density Is Associated with Individual Differences in Frontoparietal Connectivity during Working Memory.” Journal of Neuroscience 31 (40): 14284–90. https://doi.org/10.1523/JNEUROSCI.3114-11.2011.

Romer, Adrienne L., Boyu Ren, and Diego A. Pizzagalli. 2023. “Brain Structure Relations With Psychopathology Trajectories in the Adolescent Brain Cognitive Development Study.” Journal of the American Academy of Child & Adolescent Psychiatry, February. https://doi.org/10.1016/j.jaac.2023.02.002.

Romero, Jose E., Pierrick Coupé, Rémi Giraud, Vinh-Thong Ta, Vladimir Fonov, Min Tae M. Park, M. Mallar Chakravarty, Aristotle N. Voineskos, and Jose V. Manjón. 2017. “CERES: A New Cerebellum Lobule Segmentation Method.” NeuroImage 147 (February): 916–24. https://doi.org/10.1016/j.neuroimage.2016.11.003.

Samanez-Larkin, Gregory R., Joshua W. Buckholtz, Ronald L. Cowan, Neil D. Woodward, Rui Li, M. Sib Ansari, Catherine M. Arrington, et al. 2013. “A Thalamocorticostriatal Dopamine Network for Psychostimulant-Enhanced Human Cognitive Flexibility.” Biological Psychiatry, Corticostriatal Networks, Psychopathology, and Treatment, 74 (2): 99–105. https://doi.org/10.1016/j.biopsych.2012.10.032.

Sauerbrei, Willi, and Patrick Royston. 2007. “Modelling to Extract More Information from Clinical Trials Data: On Some Roles for the Bootstrap.” Statistics in Medicine 26 (27): 4989–5001. https://doi.org/10.1002/sim.2954.

Schmahmann, J. D., and J. C. Sherman. 1998. “The Cerebellar Cognitive Affective Syndrome.” Brain: A Journal of Neurology >121 (Pt 4) (April): 561–79. https://doi.org/10.1093/brain/121.4.561.

Schmahmann, Jeremy D., Xavier Guell, Catherine J. Stoodley, and Mark A. Halko. 2019. “The Theory and Neuroscience of Cerebellar Cognition.” Annual Review of Neuroscience 42: 337–64.

Stoodley, Catherine J., Jason P. MacMore, Nikos Makris, Janet C. Sherman, and Jeremy D. Schmahmann. 2016. “Location of Lesion Determines Motor vs. Cognitive Consequences in Patients with Cerebellar Stroke.” NeuroImage: Clinical 12 (February): 765–75. https://doi.org/10.1016/j.nicl.2016.10.013.

Stoodley, Catherine J., and Jeremy D. Schmahmann. 2010. “Evidence for Topographic Organization in the Cerebellum of Motor Control versus Cognitive and Affective Processing.” Cortex; a Journal Devoted to the Study of the Nervous System and Behavior 46 (7): 831–44. https://doi.org/10.1016/j.cortex.2009.11.008.

Verdejo-Garcia, Antonio, Luke Clark, Juan Verdejo-Román, Natalia Albein-Urios, José M. Martinez-Gonzalez, Blanca Gutierrez, and Carles Soriano-Mas. 2015. “Neural Substrates of Cognitive Flexibility in Cocaine and Gambling Addictions.” The British Journal of Psychiatry: The Journal of Mental Science 207 (2): 158–64. https://doi.org/10.1192/bjp.bp.114.152223.

Watson, Thomas, Nadine Becker, Richard Apps, and Matthew Jones. 2014. “Back to Front: Cerebellar Connections and Interactions with the Prefrontal Cortex.” Frontiers in Systems Neuroscience 8. https://www.frontiersin.org/articles/10.3389/fnsys.2014.00004.

Weintraub, Sandra, Sureyya S. Dikmen, Robert K. Heaton, David S. Tulsky, Philip D. Zelazo, Patricia J. Bauer, Noelle E. Carlozzi, et al. 2013. “Cognition Assessment Using the NIH Toolbox.” Neurology 80 (11 Suppl 3): S54–64. https://doi.org/10.1212/WNL.0b013e3182872ded.

Westbrook, A., and T. S. Braver. 2016. “Dopamine Does Double Duty in Motivating Cognitive Effort.” Neuron 89 (4): 695–710. https://doi.org/10.1016/j.neuron.2015.12.029.

Yao, Qun, Fanyu Tang, Yingying Wang, Yixin Yan, Lin Dong, Tong Wang, Donglin Zhu, Minjie Tian, Xingjian Lin, and Jingping Shi. 2022. “Effect of Cerebellum Stimulation on Cognitive Recovery in Patients with Alzheimer Disease: A Randomized Clinical Trial.” Brain Stimulation 15 (4): 910–20. https://doi.org/10.1016/j.brs.2022.06.004.

